# Longitudinal associations between neuropsychiatric symptoms, structural and perfusion MRI changes in older adults with Mild Cognitive Impairment

**DOI:** 10.64898/2026.10.15.26364284

**Authors:** Giada Lombardi, Cheryl Cheuk-Yan Leung, Rachel Burge, Fernando Zelaya, Sagnik Bhattacharyya, Latha Velayudhan

## Abstract

Neuropsychiatric symptoms (NPS) are highly prevalent in Mild Cognitive Impairment (MCI), conceptualized as an intermediate clinical state between normal aging and dementia. Although neuroimaging studies have reported associations between NPS and brain alterations in MCI, longitudinal evidence on NPS and their relationship with multimodal MRI measures remains limited.

We aimed to investigate (1) longitudinal changes in NPS, volumetric assessment and measurement of regional cerebral blood flow (CBF) within the medial temporal lobe (MTL); (2) associations between longitudinal changes in MRI measures and NPS; (3) whether baseline MRI measures predicted longitudinal changes in NPS.

Thirty individuals with MCI aged over 65 years were recruited, of whom eighteen followed up for a mean period of 29 ± 8.9 months.

There were significant reductions in total grey matter and white matter volumes, together with volume loss in the bilateral hippocampus, bilateral parahippocampal gyrus and right entorhinal cortex (p_FDR<_0.05) over the follow up period. Volume loss in the right parahippocampal gyrus was associated with increase in aggression (p_FDR_<0.01), and CBF increase in the right hippocampus and parahippocampal gyrus was associated with worsening in agitation (p_FDR_<0.01). Furthermore, lower baseline volume of the right entorhinal cortex was associated with increase in aggression, and lower baseline CBF in the right hippocampus and parahippocampal gyrus was associated with worsening of agitation (p_FDR_<0.05).

Our findings highlight the importance of identifying NPS in individuals with MCI and the role of structural and perfusion MRI measures as promising biomarkers for monitoring disease progression and improving risk prediction of conversion to dementia.

## Introduction

Mild Cognitive Impairment (MCI) refers to an intermediate stage between normal aging and early dementia, characterised by decline in cognitive functions such as memory, executive functioning, processing speed and reasoning [1]. In addition to cognitive symptoms, there has been increasing interest in neuropsychiatric symptoms (NPS) in people with MCI, which refer to non-cognitive disturbances involving mood, perception and behaviour [2,3]. NPS, including depression, irritability, anxiety, agitation and sleep disturbances are present in 35-85% of MCI patients [4]. Their co-occurrence has been associated with a greater functional impairment, poorer quality of life, increased caregiver burden and accelerated progression to dementia [5]. In this context, NPS may represent important targets for both early diagnosis and prevention of progression to dementia in individuals with MCI. Given their clinical significance, identifying biomarkers associated with NPS may help elucidate the biological mechanisms underlying MCI, improve risk stratification, and inform more targeted monitoring and treatment strategies. Neuroimaging offers a unique opportunity to investigate in vivo brain alterations that may contribute to the presence and progression of neuropsychiatric manifestations in MCI. Previous studies have consistently demonstrated medial temporal lobe (MTL) atrophy in affected individuals [6]. In particular, the hippocampus and entorhinal cortex have been shown to display volume reductions across the continuum from healthy ageing to MCI and Alzheimer’s disease (AD) [7, 8]. In addition to structural MRI, arterial spin labelling (ASL) MRI has emerged as a promising non-invasive technique for assessing regional cerebral perfusion and its influence on neurodegeneration.

An increasing number of studies have applied ASL to examine perfusion in MCI, showing abnormal CBF patterns across several cortical and limbic regions [9,10]. Recent evidence suggests that CBF within MTL structures may represent a potential vascular biomarker across the AD spectrum, providing complementary information to structural measures of neurodegeneration [11]. Importantly, findings regarding MTL perfusion in MCI have been considerably more heterogeneous than those observed in established AD, with reports of both hypoperfusion and hyperperfusion across hippocampal and parahippocampal regions [11]. This variability has been interpreted as reflecting dynamic vascular adaptations, including compensatory hyperperfusion during the early stages of neurodegeneration before the transition to the hypoperfusion typically observed in later disease stages [11,12].

Although neuroimaging studies have characterised structural and perfusion abnormalities in MCI, considerably less is known about how these brain alterations relate to NPS, compared with the substantial progress that has been made in understanding the neural correlates of cognitive symptoms in dementia. Emerging evidence indicates that NPS may be associated with structural and vascular brain changes, particularly within MTL, fronto-limbic and default-mode network (DMN) regions [13,14]. However, most previous work has focused either on cognitive decline and conversion to dementia, or on global measures of neuropsychiatric burden, rather than on longitudinal changes in individual NPS domains [15]. Consequently, relatively little is known about whether changes in specific NPS are associated with concurrent changes in regional brain volumes and CBF, or whether baseline MRI measures may represent predictive markers for NPS changes over time. Another important consideration concerns the assessment of NPS. Most existing studies have relied on the Neuropsychiatric Inventory (NPI) as the accepted and standard measure of NPS, which is primarily based on information provided by an informant, usually the patient’s caregiver, rather than on a clinician-integrated evaluation of symptom presentation [16]. The Neuropsychiatric Inventory-Clinician rating scale (NPI-C) was developed to provide a more comprehensive clinician-rated assessment of NPS [17]. The NPI-C is clinician rated after interviewing both patient and the carer and assesses fourteen symptom domains: delusions, hallucinations, agitation, aggression, dysphoria/depression, anxiety, elation/euphoria, apathy/indifference, disinhibition, irritability/lability, aberrant motor disturbance, sleep disorders, appetite/eating disorders and aberrant vocalisations.

Given the limited longitudinal evidence on individual NPS and neuroimaging biomarkers in MCI, the present study aimed to: (1) assess longitudinal changes of brain volumes and cerebral blood flow (CBF) in regions of MTL (hippocampus, parahippocampal gyrus and entorhinal cortex); (2) examine whether longitudinal changes in MRI measures were correlated with longitudinal changes in individual NPI-C domains; and (3) investigate whether baseline MRI measures were predictive of longitudinal changes in individual NPI-C domains. By focusing on clinician-rated NPI-C domains and longitudinal MRI data, this study seeks to improve understanding of the biological mechanisms underlying NPS progression in older individuals with MCI and to identify neuroimaging markers that may contribute to earlier detection and targeted intervention strategies.

## Methods

### Participants

Thirty participants aged over 65 years with ICD-10 diagnosis of Mild Cognitive Impairment (MCI) [18], with Clinical Dementia Rating (CDR) [19] scale score of 0.5 were recruited from memory services within the South London and Maudsley NHS Foundation Trust. Exclusion criteria comprised fulfilment of DSM-5 dementia criteria, significant neurological or psychiatric comorbidity, and severe systemic medical illness. Ethical approval was obtained from the London/Camberwell St Giles Research Ethics Committee (REC reference: 06/Q0706/50) and all participants gave written informed consent prior to the study (see Supplementary Table 1 for sociodemographic and clinical details of patients at baseline). Although this was a longitudinal study with annual assessments, the follow up was affected by the COVID-19 pandemic which made the follow up longer than a year for some participants. All thirty participants underwent a baseline visit with MRI acquisition and clinical assessments, including the standardised Mini-Mental State Examination (sMMSE) [20] and the Neuropsychiatry Inventory-Clinician version (NPI-C) for assessing neuropsychiatric symptoms. Of these, nineteen participants underwent follow-up for MRI acquisition with one of them undergoing the neuroimaging but with incomplete sMMSE [21] and NPI-C assessments. Table 1 shows the sociodemographic and clinical details of 18 patients who carried out complete baseline and follow-up MRI and clinical assessments). The average time between visits was 882.61 days (approximately 29 months). No changes in psychotropic medication were reported during the follow-up period, with the exception of one patient who was started on amitriptyline 75 mg at follow-up.

**Table 1.** Longitudinal data: summary of sociodemographic and clinical data for participants with complete MRI and clinical data at both baseline and follow-up visits. Abbreviations: sMMSE, standardized Mini-Mental State Examination; NPI-C, Neuropsychiatry Inventory­Clinician version. Unless otherwise specified, all values are expressed as mean (±standard deviation). p-values for the longitudinal changes in NPI-C domains are reported as uncorrected values and as FDR-corrected values using the Benjamini-Hochberg procedure across 14 comparisons.

| **Sociodemographic data** | *Baseline* | *Follow-up* |  |  |
| --- | --- | --- | --- | --- |
| **Sample size (n)** | 18 | 18 |  |  |
| **Age (years)** | 78.17 (±7.01) | 79.70 (±6.77) |  |  |
| **Sex (Females/Males)** | 12/6 | 12/6 |  |  |
| **Education (years)** | 14.0 (±4.79) | 14.0 (±4.79) |  |  |
| **Clinical measures (total scores)** | *Baseline* | *Follow-up* | *p-value* |  |
| **sMMSE** | 26.00 (±3.48) | 26.23 (±2.41) | 0.95 |  |
| **NPI-C** | 26.94 (±22.89) | 18.11 (±14.98) | 0.17 |  |
| **NPI-C items** | *Baseline* | *Follow-up* | *p-value* |  |
|  |  |  | *uncorr* | *corr* |
| **Delusions** | 0.17 (±0.38) | 0.39 (±1.42) | 0.45 | 0.70 |
| **Hallucinations** | 0.33 (±0.84) | 0.17 (± 0.38) | 0.33 | 0.66 |
| **Agitation** | 5.22 (±5.31) | 2.0 (± 2.59) | 0.002 | 0.03 |
| **Aggression** | 1.17 (±1.65) | 1.0 (± 3.07) | 0.03 | 0.66 |
| **Dysphoria** | 4.0 (±6.43) | 2.78 (± 3.84) | 0.65 | 0.81 |
| **Anxiety** | 2.78 (±4.85) | 2.39 (± 5.10) | 0.69 | 0.81 |
| **Elation/Euphoria** | 0.06 (±0.24) | 0.22 (± 0.55) | 0.27 | 0.63 |
| **Apathy/Indifference** | 4.22 (±8.91) | 3.67 (± 10.31) | 0.23 | 0.63 |
| **Disinhibition** | 0.72 (±1.18) | 0.72 (± 1.13) | 0.89 | 0.90 |
| **Irritability/Lability** | 2.22 (±4.05) | 0.94 (± 1.59) | 0.21 | 0.63 |
| **Aberrant motor disturbance** | 0.61 (±1.09) | 0.89 (± 1.41) | 0.42 | 0.70 |
| **Sleep disorders** | 4.22 (±5.04) | 2.94 (± 3.40) | 0.30 | 0.70 |
| **Appetite/Eating disorder** | 0.89 (±1.41) | 0.78 (± 0.94) | 0.67 | 0.85 |
| **Aberrant vocalization** | 0.33 (±1.19) | 1.67 (± 3.88) | 0.86 | 0.90 |

### MRI acquisition

MRI data were acquired using a General Electric (GE) Discovery MR750 3TDMRID scanner with a 12-channel radiofrequency head coil at the Centre for Neuroimaging Sciences, King’s College London. Structural images were obtained using a □ sagittal 3D T1-weighted accelerated magnetisation-prepared rapid gradient-echo sequence (SAG ACC MPRAGE) (TR = 7.31 ms, TE = 3.02 ms, 256 x 256 matrix, 196 slices, voxel size 1.2 x 1.05 x 1.05 mm**^3^**); Cerebral perfusion images were acquired using a 3D pseudo-continuous Arterial Spin Labelling (3D-pCASL) sequence, using a 1800 ms pseudo-continuous RF labelling pulse, a 2025 ms post-labelling delay, and a 3D ‘stack of spirals’ readout that provided 60 axial slice locations (3 mm thickness) and an in-plane resolution of 1.8mm, after transformation to a rectangular matrix (TR = 5.18 s, TE = 0.011 s, flip angle = 111°). Four ‘control-labelled’ pairs were acquired, using four ‘background suppression’ pulses to minimise static signal. A proton density (PD) image volume with the same parameters was acquired within the same sequence parameters in the readout, to use as a reference to compute the CBF maps in conventional physiological units (mL blood/100 g tissue/minute).

### MRI preprocessing and analysis

Structural T1-weighted MRI images were processed using FastSurfer, a deep-learning–based pipeline for cortical surface reconstruction and volumetric segmentation [22]. First, skull stripping was performed using HD-BET [23]. The resulting images were then processed with the FastSurfer pipeline using the *run_fastsurfer.sh* workflow, which performs cortical and subcortical segmentation based on a convolutional neural network. Segmentation outputs were aligned to the subject native T1 space using affine registration implemented in ANTs (Advanced Normalization Tools; https://github.com/ANTsX/ANTs). Additionally, the Computational Anatomy Toolbox (CAT12) running on SPM [24] was used to extract Total intracranial volume (TIV), as well as Grey Matter (GM), White Matter (WM) and Cerebrospinal fluid (CSF) volumes.

Preprocessing of the ASL images was carried out by using an open-source data processing toolbox (ASAP) based on MATLAB [25]. Cerebral blood flow (CBF) maps were calculated using the currently recommended method [26], using the proton density (PD) images as reference. The resulting CBF maps were then rigidly co-registered to the individual T1-weighted anatomical images using the PD image to improve co-registration accuracy. The T1 weighted images were subsequently normalized to the MNI standard space and the transformation matrix was applied to the CBF maps. Finally, the maps were spatially smoothed using a Gaussian kernel (FWHM of 6 mm).

Following preprocessing, region-of-interest (ROI)-based analysis was performed to extract regional volumes and mean regional CBF values. Particularly, we considered regions of the Medial Temporal Lobe (MTL) including right/left hippocampus, right/left parahippocampal gyrus and right/left entorhinal cortex, based on previous evidence showing early structural and perfusion alterations within these regions in MCI and AD [7–12].

Structural volumes of the selected ROIs were extracted directly from the FastSurfer segmentation outputs. The FastSurfer Desikan–Killiany–Tourville (DKT) atlas parcellation was also used for CBF extraction in the same ROIs.

### Statistical analysis

Statistical analyses were performed using IBM SPSS Statistics (version 31.0.1.0). Normality of data distributions was assessed using the Shapiro-Wilk test and visual inspection of Q-Q plots. Paired comparisons between baseline and follow-up visits were performed for each clinical variable (sMMSE and NPI-C) and global MRI measures, including GM, WM and TIV. Paired t-tests were applied for normally distributed variables, whereas the Wilcoxon signed-rank test was used for variables showing non-normal distributions. The NPI-C total score was analysed as a single outcome measure, whereas longitudinal changes in the individual NPI-C domains were analysed separately. To account for multiple testing across NPI-C domains (14 comparisons), p-values from the individual domain analyses were adjusted using the Benjamini–Hochberg False Discovery Rate (FDR) procedure.

Longitudinal changes in regional brain volumes and cerebral blood flow (CBF) were assessed using the same approach, with p-values adjusted using FDR correction applied to account for multiple comparisons across regions of interest (ROIs) within MTL (6 comparisons).

Exploratory analyses were subsequently conducted for NPI-C domains showing evidence of longitudinal change (Δ = follow-up - baseline) at the uncorrected level (p < 0.05). Associations between longitudinal MRI changes and longitudinal NPS changes were examined using Spearman rank correlations. In addition, exploratory linear regression analyses were performed to investigate whether baseline MRI measures were associated with longitudinal changes in NPS. Baseline regional volumes and CBF values were entered as independent variables and change scores for the selected NPI-C domains as dependent variables.

Follow-up interval was included as a covariate, and FDR correction was applied to account for multiple comparisons. In addition, to account for potential confounding effects of vascular comorbidities on cerebral perfusion, diabetes, hypertension, cerebrovascular accident (CVA) and myocardial infarction (MI) were included as covariates (0 = absent, 1 = present) in analyses involving CBF measures.

## Results

### Baseline

Supplementary Table 1 reports sociodemographic and clinical data for all individuals with MCI who completed baseline assessments (n=30).

### Longitudinal changes

For MCI patients, longitudinal analysis of clinical measures using Wilcoxon signed-rank tests did not reveal a significant difference between baseline and follow-up for both total scores of sMMSE (W=54.0, p=0.95) and NPI-C (W=137.5, p=0.071). Among the individual NPI-C domains, agitation showed a significant longitudinal change (reduction) that survived FDR correction (p_FDR_=0.042), while aggression showed evidence of longitudinal change (reduction) at the uncorrected level only (p=0.03).

Results of paired t-tests of global structural MRI measures revealed a significant reduction of both GM (t(18)=2.30, p=0.034, Figure 1 A1) and WM (t(18)=3.32, p=0.004, Figure 1 A2) volumes, and a reduction of TIV close to significance (t(18)=1.94, p=0.07).

**Figure 1.**
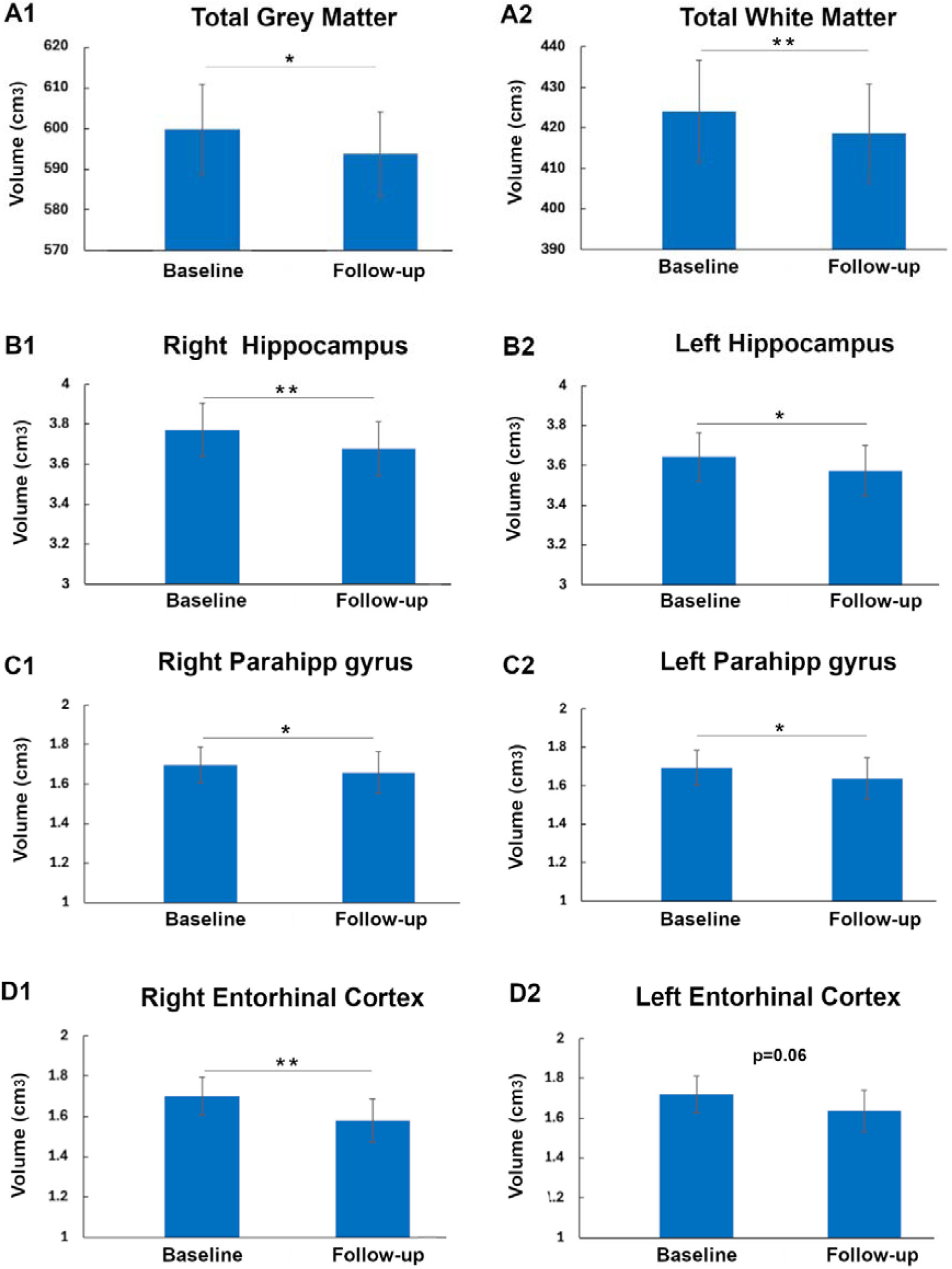
Longitudinal MRI changes of total GM (A1) and WM (A2) volumes, right (B1) and left (B2) hippocampus, right (C1) and left (C2) parahippocampal gyrus, right (D1) and left (D2) entorhinal cortex. Vertical bars represent standard error of the mean (SEM). Horizontal bars represent statistical significance (* p 0.05, ** p 0.01).

**Figure 2.**
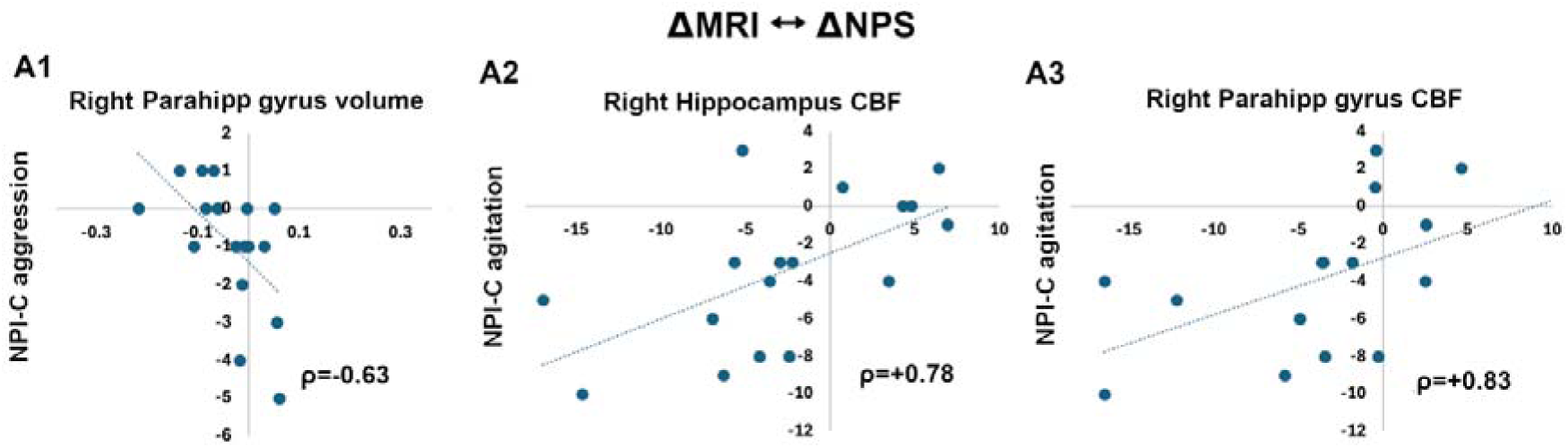
Results of correlations between longitudinal changes (A = follow-up - baseline) in MRI measures and neuropsychiatric symptoms (AMRI ↔ ANPS). Scatter plots show the relationships between changes in (A1) right parahippocampal gyrus volume and NPI-C aggression symptoms, (A2) right hippocampal CBF and NPI-C agitation symptoms, (A3) right parahippocampal gyrus CBF and NPI-C agitation symptoms. Spearman rank correlation coefficients (p) are reported for each analysis, with p<0 indicating negative associations and p>0 indicating positive associations.

Results of paired t-tests of regional volumes revealed a significant volume reduction of the right hippocampus (t(18)=3.81, p=0.001, p_FDR_<0.01, Figure 1 B1), left hippocampus (t(18)=2.22, p=0.04, p_FDR_=0.05, Figure 1 B2), right parahippocampal gyrus (t(18)=2.24, p=0.039, p_FDR_=0.048, Figure 1 C1), left parahippocampal gyrus (t(18)=3.07, p=0.007, p_FDR_=0.014, Figure 1 C2) and right entorhinal cortex (t(18)=4.3, p<0.001, p_FDR_<0.01, Figure 1 D1). The left entorhinal cortex showed a trend toward volume reduction (t(18)=2.00, p=0.06), which did not survive FDR correction (Figure 1 D2). No significant CBF changes surviving FDR correction were detected.

Subsequent correlation and regression analyses were restricted to agitation and aggression, as these were the only NPI-C domains showing evidence of longitudinal change over time (agitation surviving FDR correction and aggression at the uncorrected level).

### Correlations between longitudinal changes of MRI and NPS

For structural MRI measures, a significant negative correlation surviving FDR correction was observed between change in right parahippocampal gyrus volume and change in NPI-C aggression scores (ρ=-0.632, pFDR<0.01, Figure 2 A1), indicating that volume loss was associated with increase in aggression symptoms over the follow up period.

For ASL measures, significant positive correlations surviving FDR correction were observed between NPI-C agitation scores change and CBF change in the right hippocampus (p=0.78, Pfdr <0.01, Figure 2 A2) and right parahippocampal gyrus (p=0.83, pfdr <0.01, Figure 2 A3), indicating that CBF increase in these regions was associated with increase in agitation domain score over time.

### Baseline MRI measures predict longitudinal NPS change

For structural MRI measures, baseline volume of the right entorhinal cortex (0=-2.4, R=0.63, p_FDR_<0.05) was associated with NPI-C aggression score changes (Figure 3 A1).

**Figure 3.**
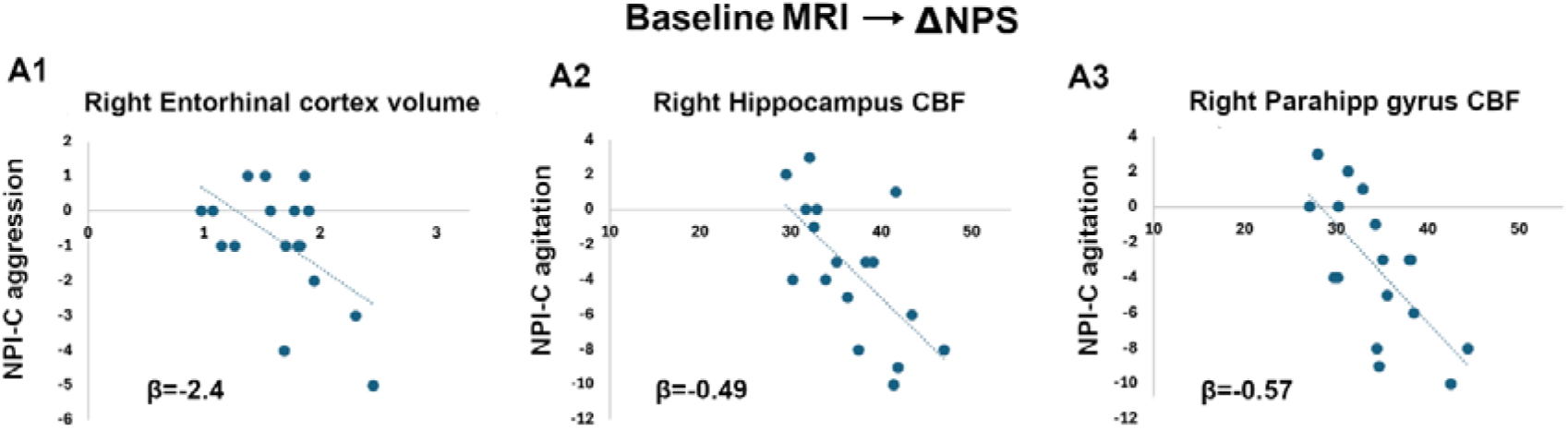
Regression analysis results with baseline MRI measures as independent variables and longitudinal changes of neuropsychiatric symptoms as dependent variables (Baseline MRI -> ANPS). (A1) Baseline volumes of the right entorhinal cortex associated with changes in NPI-C aggression symptoms; (A2) baseline CBF of the right hippocampus and (A3) right parahippocampal gyrus associated with changes in NPI-C agitation symptoms. Regression coefficients (β) are reported for each analysis, with β<0 indicating negative associations.

For ASL measures, baseline CBF values in the right hippocampus (p=-0.49, R=0.75, Pfdr<0.05, Figure 3 A2), and right parahippocampal gyrus (0=-0.57, R=0.78, Pfdr<0.05, Figure 3 A3) were associated with NPI-C agitation score changes.

## Discussion

The present study investigated longitudinal changes in neuropsychiatric symptoms (NPS), regional brain volumes and cerebral blood flow (CBF) and their relationship in older adults with MCI.

Consistent with previous research, longitudinal analysis demonstrated significant reductions in global grey matter and white matter volumes [27], together with significant volume loss in regions of the MTL, including the hippocampus, parahippocampal gyrus and entorhinal cortex. Histopathological evidence indicates that the hippocampus and entorhinal cortex are among the earliest regions affected in MCI, with patients showing smaller volumes compared with cognitively healthy older adults [28]. Previous MRI studies have further demonstrated progressive volume reductions in these structures across the continuum from normal ageing to MCI and then to AD, supporting their role as key markers of early neurodegeneration [7; 29] including the AddNeuroMed study [30]. Furthermore, previous longitudinal studies showed that smaller hippocampal and entorhinal cortex volumes are associated with an increased likelihood of progression from MCI to AD [31, 32], while accelerated atrophy rates in these structures have been linked to subsequent clinical deterioration [33]. Together, these findings support the view that hippocampal and entorhinal cortex atrophy may represent an important neuroimaging marker of disease progression during the prodromal stages of dementia.

Despite evidence of progressive structural brain changes, the total score of NPI-C did not differ significantly between visits. Although most NPI-C domains showed lower scores at follow-up, only agitation showed a significant reduction after multiple comparisons, while aggression showed evidence of a longitudinal reduction at the uncorrected level only. Several factors may have contributed to this overall pattern of improvement. Firstly, some of the recruitment and baseline assessments were conducted during the COVID-19 pandemic, a period associated with increased social isolation and psychological distress, which may have transiently influenced NPS and potentially contributed to higher symptom burden at baseline. Secondly, NPI-C assessments were carried out by different clinicians across visits, which may have introduced some inter-rater variability in symptoms ratings despite the established reliability and validity of the NPI-C [17]. Nevertheless, the variability observed across individual NPI-C domains suggests that NPS may follow distinct trajectories in MCI, rather than changing uniformly over time. This further highlights the limitation of relying solely on summary NPS measures and supports the importance of investigating individual domains separately.

Greater volume loss within the right parahippocampal gyrus was found to be associated with increase in aggression score change over the follow up period. Given that change scores were calculated as follow-up minus baseline values and that higher NPI-C scores indicate greater symptom severity, this negative correlation suggests that progressive atrophy (ΔVolume<0) within MTL is associated with worsening of agitation symptoms (ΔNPI-C>0). Emerging evidence suggests that NPS are associated with structural abnormalities within medial temporal and limbic regions across the AD spectrum [34]. Of note, as key component of the MTL, the parahippocampal gyrus is involved not only in memory processing but also in emotional and behavioural regulation [35]. Progressive atrophy within this region may therefore contribute to disruption of neural networks supporting affective and behavioural functioning, providing a plausible neurobiological explanation for the observed worsening of aggression score changes. Moreover, significant positive correlations were observed between changes in right hippocampal and parahippocampal CBF and changes in agitation scores, meaning that an increase of CBF within these regions (ΔCBF>0) was associated with agitation worsening (ΔNPI-C>0). Increased perfusion in the hippocampus has previously been reported in AD as reflecting compensatory vascular responses, increased neuronal activity or inflammatory processes [12]. NPS in AD have also been linked to pathological changes involving the hippocampus, amygdala and other limbic structures [36], while recent models of agitation emphasise dysregulation of neural circuits involved in emotional reactivity and behavioural control [37]. These data suggest that agitation may be particularly associated with alterations in MTL perfusion, supporting the notion that perfusion abnormalities may contribute to neuropsychiatric manifestations alongside structural changes.

Baseline volume of the right entorhinal cortex was found to be associated with subsequent changes in aggression scores over the follow up period indicating that lower baseline volumes predicted worsening of aggression symptoms over time. This finding is notable in light of the longitudinal structural changes observed within the MTL including significant atrophy of the right entorhinal cortex. In addition, while CBF increase in the right hippocampus and parahippocampal gyrus was associated with increase of agitation symptoms over time, lower baseline CBF within these regions predicted subsequent worsening of agitation, supporting a role for hippocampal and parahippocampal perfusion in agitation trajectories, suggesting that alterations within this system may contribute to both vulnerability to, and progression of, agitation symptoms in MCI.

The present study has several strengths. Firstly, the longitudinal design enabled the examination of symptoms and brain changes over time. Secondly, a multimodal MRI approach provided complementary information regarding brain volume and perfusion alterations in MCI. Thirdly, NPS were assessed using the clinician-rated NPI-C, allowing the investigation of individual symptom domains rather than caregiver-reported NPS scores. This approach may be particularly valuable in MCI, where they still have capacity to share their symptoms and experiences and also where the neuropsychiatric manifestations are often heterogeneous and may follow different trajectories over time [38]. However, limitations of the study should also be acknowledged. Firstly, our results should be considered in light of the modest sample size and need further confirmation in larger cohorts of patients. Secondly, given the relatively small sample size, analyses were restricted to regions of interest selected on the basis of previous evidence in MCI. While this hypothesis-driven approach reduced the burden of multiple comparisons, it may have overlooked additional brain regions involved in NPS change. Thirdly, it would be useful to have comparison group of age- and sex-matched healthy controls. Although age- and sex-matched healthy control (HC) data from our site for the AddNeuroMed study [31, 39] were available for comparison of baseline sMMSE scores and structural neuroimaging measures, these HC participants did not undergo NPI assessments or ASL imaging. Therefore, longitudinal comparisons with HC were not possible. Future studies including longitudinally followed HC would help clarify the specificity of these MRI changes to MCI-related disease processes. Finally, although several baseline MRI measures were associated with subsequent changes in NPS, the lack of data on dementia conversion prevented assessment of their prognostic value for later clinical progression.

In conclusion, this study showed that brain atrophy and cerebral perfusion within MTL are associated with changes in individual NPS domains, and that both structural and perfusion MRI measures were associated with symptoms progression. As MCI may represent a critical window for intervention before the onset of dementia, identifying biomarkers capable of detecting early neurobiological changes and improving risk stratification is of considerable clinical importance. The present findings suggest that multimodal MRI measures may provide complementary information regarding the neural substrates underlying NPS progression and may contribute to the early identification of individuals at greater risk of adverse clinical outcomes.

## Data Availability

All data produced in the present study are available upon reasonable request to the authors

## Author contributions

GL performed data analysis and wrote the first draft of the manuscript; CCYL, RB. and LV contributed to data collection and data cleaning; FZ contributed to data analysis and data interpretation; SB contributed to data interpretation. LV conceptualized the study, supervised the development of the study and contributed to data interpretation. All co-authors read the manuscript and approved its final version.

