## Supplementary material for "Longitudinal associations between neuropsychiatric symptoms, structural and perfusion MRI changes in older adults with Mild Cognitive Impairment"

| **Sociodemographic data** | *Baseline* |
| --- | --- |
| **Sample size** (n)  **Age** (years)  **Sex** (Females/Males)  **Education** (years) | 30  78.10 (±6.73)  18/12  13.27 (±4.19) |
| **Clinical measures (total scores)** | *Baseline* |
| **sMMSE**  **NPI-C** | 25.87 (±3.04)  32.10 (±28.80) |
| **NPI-C items** | *Baseline* |
| **Delusions**  **Hallucinations**  **Agitation**  **Aggression**  **Dysphoria**  **Anxiety**  **Elation/Euphoria**  **Apathy/Indifference**  **Disinhibition**  **Irritability/Lability**  **Aberrant motor disturbance**  **Sleep disorders**  **Appetite/Eating disorder**  **Aberrant vocalization** | 0.27 (±0.64)  0.27 (±0.69)  5.20 (±5.26)  1.70 (±2.28)  4.57 (±5.92)  2.67 (±4.26)  0.17 (±0.75)  4.60 (±8.70)  1.57 (±2.94)  4.47 (±7.04)  0.50 (±1.01)  4.50 (±4.73)  1.17 (±2.15)  0.47 (±1.17) |

**Supplementary Table 1. Summary of sociodemographic and clinical data for all participants who completed baseline assessments. Abbreviations: MMSE, standardized Mini-Mental State Examination; NPI-C, Neuropsychiatry Inventory-Clinician version. Unless otherwise specified, all values are expressed as mean (±standard deviation).**

**Exploratory whole-brain analysis**

Exploratory whole-brain voxel-wise analyses were performed on structural MRI and Arterial Spin Labelling (ASL)-derived cerebral blood flow (CBF) maps. For structural MRI analyses, voxel-based morphometry (VBM) was performed on normalized and smoothed grey matter images (smwp1 files) generated using the CAT12 toolbox. Statistical analyses were performed using FSL randomise with non-parametric permutation testing (5000 permutations). Statistical significance was assessed using threshold-free cluster enhancement (TFCE) with family-wise error correction (p_FWE_<0.05).

For ASL analyses, preprocessed CBF images were analysed using the same voxel-wise non-parametric permutation framework, with statistical significance assessed using TFCE and family-wise error correction (p_FWE_<0.05).

Significant clusters showing longitudinal changes in grey matter volume between baseline and follow-up identified by the whole-brain structural analysis are reported in the Supplementary Table 2. No significant voxel-wise longitudinal changes of CBF were identified.

| **Cluster** \| | **Size (n voxels)** | **Peak coordinates** | **Anatomical label** |
| --- | --- | --- | --- |
| 1 | 530 | x=-31.5, y=-6, z=-24 | Parahippocampal gyrus, anterior division |
| 2 | 109 | x=4.5, y=-10.5, z=13.5 | Right Thalamus |
| 3 | 77 | x=37.5, y=-51, z=-31.5 | Right Cerebellar Crus I/ Right Cerebellar VI |
| 4 | 61 | x=-30, y=-42, z=-13.5 | Temporal fusiform cortex/  Parahippocampal gyrus, posterior division |
| 5 | 61 | x=9, y=-25.5, z=16.5 | Right Thalamus |
| 6 | 26 | x=-49.5, y=-13.5, z=-3 | Superior Temporal gyrus |
| 7 | 8 | x=0, y=37.5, z=25.5 | Paracingulate gyrus |

**Supplementary Table 2**. **Significant TFCE-corrected clusters showing longitudinal grey matter volume reduction between baseline and follow-up.** **Peak coordinates are given in MNI space, and anatomical labels were assigned using the Harvard–Oxford cortical and subcortical atlases and the cerebellar atlas in MNI space.**
